# Protocol for a Magnetic Resonance Imaging (MRI) Study of Participants in the Fever Randomized Controlled Trial: Does fever control prevent brain injury in malaria?

**DOI:** 10.1101/2023.11.10.23298374

**Authors:** Moses B. Chilombe, Karl B. Seydel, Colleen Hammond, Suzanna Mwanza, Archana A. Patel, Frank Lungu, Somwe wa Somwe, Sam Kampondeni, Michael J. Potchen, Michael P. McDermott, Gretchen L. Birbeck

## Abstract

**Background:** Despite eradication efforts, ∼135,000 African children sustained brain injuries as a result of central nervous system (CNS) malaria in 2021. Newer antimalarial medications rapidly clear peripheral parasitemia and improve survival, but mortality remains high with no associated decline in post-malaria neurologic injury. A randomized controlled trial of aggressive antipyretic therapy with acetaminophen and ibuprofen (Fever RCT) for malarial fevers being conducted in Malawi and Zambia began enrollment in 2019. We propose to use neuroimaging in the context of the RCT to further evaluate neuroprotective effects of aggressive antipyretic therapy.

**Methods:** This observational magnetic resonance imaging (MRI) ancillary study will obtain neuroimaging and neurodevelopmental and behavioral outcomes in children previously enrolled in the Fever RCT at 1- and 12-months post discharge. Analysis will compare the odds of any brain injury between the aggressive antipyretic therapy and usual care groups based upon MRI structural abnormalities. For children unable to undergo imaging without deep sedation, neurodevelopmental and behavioral outcomes will be used to identify brain injury.

**Discussion:** Neuroimaging is a well-established, valid proxy for neurological outcomes after brain injury in pediatric CNS malaria. This MRI ancillary study will add value to the Fever RCT by determining if treatment with aggressive antipyretic therapy is neuroprotective in CNS malaria. It may also help elucidate the underlying mechanism(s) of neuroprotection and expand upon FEVER RCT safety assessments.

## Background

Central nervous system (CNS) malaria, meaning *P. falciparum* parasitemia with neurologic symptoms including impaired consciousness and/or seizures,^1–4^ continues to be a major cause of neuro-disabilities in African children. Brain injury with subsequent neurologic sequelae occurs in one-third of children who survive CNS malaria. Newer antimalarial medications rapidly clear peripheral parasitemia and improve survival, but mortality remains high with no documented decline in post-malaria neurologic injury.^5^ Common sequelae include epilepsy, gross motor deficits, blindness, cognitive impairment, and behavioral disorders.^1–4, 6–8^

In the Blantyre Malaria Project Epilepsy Study (BMPES), a higher maximum temperature (Tmax) during the acute infection was associated with a greater risk of post-malaria neurologic sequelae. Children who developed epilepsy had a mean Tmax of 39.4°C vs. 38.5°C in those who did not develop epilepsy (absolute difference of 0.9°C; p=0.01). In addition, those with subsequent behavioral disorders had a higher Tmax than those who did not (39.2°C vs. 38.7°C; absolute difference 0.5°C; p=0.04).^1^ These data are consistent with the substantial body of evidence that, within the peri-insult period, fevers contribute to secondary neurologic injury across a wide range of conditions including stroke, meningitis, anoxia, hypoxia, and trauma.^9–13^ In the United States (US), induced hypothermia is now standard-of-care for post-anoxic^14^ and neonatal hypoxic-ischemic encephalopathy^15–18^ and fever prevention is an important aspect of neurocritical care.^19–22^ In contrast to this aggressive and proactive approach to fever control, the World Health Organization (WHO) malaria care guidelines recommend only modest measures for fever control and these are to be used only in response to a significant febrile event.

Specifically, WHO malaria guidelines recommend treating a core temperature of ≥ 38.5°C with acetaminophen 15 mg/kg (no loading dose) and explicitly recommend against the use of other non-steroidal medications.^23^ Malaria-induced fevers can be extremely high and there is no evidence to date that a single antipyretic reduces malaria fevers. A controlled trial of acetaminophen vs. placebo for uncomplicated malaria found no differences in mean temperature, but 4% of children receiving placebo required admission for febrile seizures.^24^ In BMPES, WHO malaria care guidelines were followed and 15% of children experienced at least one temperature of ≥ 40.0°C. The age-related susceptibility to pediatric malaria substantially overlaps with the age-related phenomena of febrile seizures. Malarial seizures are often complex, multifocal, and prolonged with status epilepticus being common.^1, 7, 25, 26^ No malaria studies to date have evaluated the impact of interventions using two antipyretics.

In 2019, a randomized controlled trial of aggressive antipyretic therapy for CNS malaria (Fever RCT) launched in Malawi and Zambia.^27^ This RCT compares prophylactic, scheduled acetaminophen, including an initial loading dose of 30 mg/kg, plus ibuprofen for 72 hours after presentation with CNS malaria to usual care based upon WHO guidelines over that same period. The primary outcome of the RCT is Tmax. This observational MRI ancillary study will determine whether the aggressive antipyretic therapy used in the Fever RCT resulted in fewer brain injuries based upon neuroimaging or neurodevelopment and behavioral outcomes where safe imaging is not possible.

### Rationale

While a higher Tmax is associated with adverse neurologic outcomes in CNS malaria, causality is not established. CNS malaria is a highly inflammatory process^28, 29^ and inflammation-mediated injury is a likely pathway for malaria-associated brain injury in this population, particularly for epilepsy development. The aggressive antipyretic (AA) regimen used in the Fever RCT may offer anti-inflammatory benefits in addition to fever-reduction. Neuroimaging of the Fever RCT participants will facilitate identification of possible neuroprotective benefits from AA treatment including anti-inflammatory benefits that might not be evident based upon acute temperature measures. Previous research among cerebral malaria survivors in Blantyre has shown that 53% suffer from cognitive and/or behavioral impairments at one year and these impairments were associated with structural brain lesions on MRI including severe atrophy and multifocal abnormalities.^30^

This study will also expand the safety assessments of the AA intervention. Ibuprofen may cause bleeding due to its antiplatelet effects and is also nephrotoxic. Though frank CNS bleeds are not associated with pediatric cerebral malaria,^31^ end vessel micro-hemorrhages are seen post mortem^32, 33^ and have been identified on brain MRIs.^34, 35^ MRI gradient recalled echo (GRE) sequences available on standard MR equipment are sensitive to ferromagnetic substances and will detect micro-hemorrhages. In Zambia where standard imaging is available, we will compare the presence and characteristics of any CNS bleeding in the AA vs usual care (UC) groups.

Newly published data emerged after the Fever RCT commenced that shows a link between cerebral malaria, acute kidney injury and the development chronic kidney disease (CKD),^36^ so the development of CKD after CNS malaria will also be compared using creatinine and the urine albumin:creatinine ratio (ACR) assessed at least 6-months post-recovery.

### Main Objectives

This prospective observational study will seek consent from parents of children previously enrolled in the Fever RCT (Aggressive Antipyretics in CNS Malaria: A Randomized Controlled Trial Assessing Antipyretic Efficacy and Parasite Clearance; National Institutes of Health Grant R01NS102176; ClinicalTrials.gov NCT03399318) to obtain neuroimaging and neurodevelopmental & behavioral outcomes data on their child. The imaging and evaluations for this observational study will occur after the child has recovered from the acute malaria infection and has otherwise completed the Fever RCT study. For some, consent will be sought months after the RCT-related activities for the child are completed. As such, this is an independent observational study.

### Specific Aims

#### Aim 1

To compare the prevalence of brain injury in the AA vs UC groups of the Fever RCT. Children will undergo brain MRIs and neurodevelopmental & behavioral evaluations at 1 and 12 months after their acute malaria illness or as soon as possible among those children enrolled in the Fever RCT prior to the initiation of this MRI ancillary study. Two radiologists, blinded to treatment allocation, will independently review images and capture data using NeuroInterp.^37^ For children who are unable to undergo imaging without deep sedation, neurodevelopmental & behavioral evaluations will be used to identify those with brain injury.

We hypothesize that children who receive AA therapy during CNS malaria will have lower odds of brain injury than those receiving UC.

#### Aim 2

To compare the prevalence of specific structural injuries among children who received AA therapy vs. UC therapy in the Fever RCT. Specifically, we will compare

a. Overall atrophy by brain volume scoring^38^ (atrophy being a brain volume score of 1-2 vs ≥3)
b. Gliosis by Fazekas score^39^ (0, 1, 2 or 3)
c. The presence of regional gliosis or atrophy in the following regions

1. Cortical (present/absent)
2. Deep grey (present/absent)
3. Corpus callosum (present/absent)
4. Posterior fossa (present/absent)

*A priori* pathophysiologic attributions are as follows:

– If focal cortical abnormalities are lower for the AA treatment group, we will conclude that the mechanism(s) included seizure control/prevention
– If abnormalities are less common in the deep grey regions for the AA treatment group, we will attribute this to benefits of reduced sequestration in the deep vascular beds
– If there is less diffuse atrophy or overall gliosis for the AA treatment group, we will attribute this to reductions in increased intracranial pressure

#### Aim 3: Safety assessments

a. In Zambia where standard imaging with GRE sequences is available, we will compare the prevalence of blood products in the AA vs UC groups. If present, blood products will be further characterized by age of blood relative to participation in the Fever RCT and blood volume based upon number and size of foci.
b. The prevalence of CKD based upon blood creatinine and urine ACRs will be compared in the AA vs UC groups

## Methods

All patient-contact will occur in Zambia and Malawi. All data transmitted or conveyed beyond the country of enrollment will be de-identified.

### Study sites

- University Teaching Hospitals-Children’s Hospital, Lusaka,
- Chipata Central Hospital, Chipata, Zambia
- Queen Elizabeth Central Hospital, Blantyre, Malawi

Study sites without patient contact or access to data with private health information include:

- University of Rochester, Rochester, NY, USA
- Michigan State University, East Lansing, MI, USA

### Study design

Consented participants from the Fever RCT will undergo (1) a non-contrast brain MRI, (2) neurodevelopmental & behavioral assessment, and (3) kidney function assessments with blood creatinine and urine ACR studies at 1 and 12 months post recovery, or as soon as possible if their Fever RCT admission occurred prior to the launch of this observational study. Based upon planned enrollment for the RCT (284), and considering anticipated mortality and loss to follow-up, expected enrollment is 184 total.

### Subject population

All children enrolled in the Fever RCT who survive to discharge are eligible for enrollment. Inclusion and exclusion criteria for the Fever RCT are outlined below.

#### Eligibility criteria

### Inclusion Criteria for RCT

- Age 2-11 years (24 to 132 months)
- Evidence of *P. falciparum* malaria infection by peripheral blood smear or rapid diagnostic test
- CNS symptoms associated with malaria. This includes the most severe manifestation, that being cerebral malaria (CM) with impaired consciousness via Blantyre Coma Score (BCS) ≤2 in children under 5 years or a Glasgow Coma score (GCS) ≤10 in children ≥5 years; or less severe disease with complicated seizure(s), meaning prolonged (>15 minutes), focal or multiple; or other evidence of impaired consciousness (confusion, delirium) without frank coma (BCS>2, GCS=11-14)

### Exclusion Criteria for RCT

- Circulatory failure (cold extremities, capillary refill > 3 seconds, sunken eyes, decreased skin turgor)
- Vomiting in the past 2 hours
- Serum Cr > 1.2 mg/dL
- A history of liver disease
- Jaundice or a total bilirubin of >3.0mg/dL
- A history of gastric ulcers or gastrointestinal bleeding
- A history of thrombocytopenia or other primary hematologic disorder
- Petechiae or other clinical indications of bleeding abnormalities
- A known allergy to ibuprofen, acetaminophen, aspirin, or any non-steroidal medication
- Any contraindication for nasogastric tube placement and/or delivery of enteral medications

### Recruitment Methods

For children enrolled in the Fever RCT before this MRI ancillary study was commenced, consent for future contact regarding possible research participation in subsequent relevant studies was sought from the parents/guardians. This process was approved by the appropriate ethical and institutional review boards. A list of those who consented by study identifiers (IDs) will be cross-referenced to the contact details including address and cell phone numbers maintained in the study pharmacies. These parents/guardians will be contacted to seek consent for enrollment in this observational study. If the parent/guardian is willing to consider enrollment, they will be invited to come to the hospital for a full discussion and the consent process detailed below.

Children enrolled in the Fever RCT after this MRI ancillary study is launched will be identified as potential participants when they have been stabilized and regained consciousness. Study nurses working on the ward where the children are admitted will then approach the parent/guardian for consent prior to discharge.

### Consent Process

For children whose parents consented to be contacted regarding future studies, phone calls will be placed and a brief overview of the study will be provided. Consent and assent forms will be developed in English and the applicable local language. Forward- and back-translation will be undertaken by expert local staff with extensive experience in this regard. Staff obtaining consent are fluent in the local languages and expert in conducting the informed consent discussion. All have Human Subjects Protection and Good Clinical Practice certifications. The consent process will be conducted in the preferred language of the parent/guardian. All parents/guardians who consent will be provided with signed copies of the consent forms. For those parents/guardians who are unable to sign due to literacy limitations, a thumb print will be obtained and someone who is not associated with the study but is literate in the parent’s/guardian’s preferred language will be present during the consent process and will sign as a witness. At the time of the return visit for evaluations, for children 7 years or older, if in the parent’s/guardian’s assessment the child has the capacity to comprehend the nature of the request, assent will also be sought.

### Study Procedures

#### Imaging

In Zambia, a 0.35T GE MRI will be used to obtain T1, T2 FRFSE, FLAIR, diffusion-weighted imaging, GRE, GRE T2*, proton density, STIR and 2-D and volumetric imaging. In Malawi, a Hyperfine MRI adjacent to the research ward will be used and provides please insert. MRI images will be reviewed by the technician at the time of the scanning and any findings of concern that might warrant gadolinium administration will be reviewed by one of the study radiologists in real time to allow for gadolinium administration when the child is initially imaged. However, if on review of an MRI about which the technical staff had no concerns the radiologist feels gadolinium is warranted for clinical care, the patient’s family will be notified and all the same financial and logistical support will be provided for this repeat image. Two radiologists, blinded to treatment allocation and neurologic status, will independently review images and capture data using NeuroInterp. Disagreements will be reconciled first by consensus review and if reviewers still disagree, a third reader will make the final determination.

#### Neurodevelopmental & Behavioral

At the 1- and 12-month evaluations, age-appropriate assessments will be undertaken by dedicated local staff fully trained to deliver these measures. Data entry will be on tablets at the time of assessment. See Table 1 for details regarding instruments and criteria for a child to be categorized as brain injured.

**Table 1:**
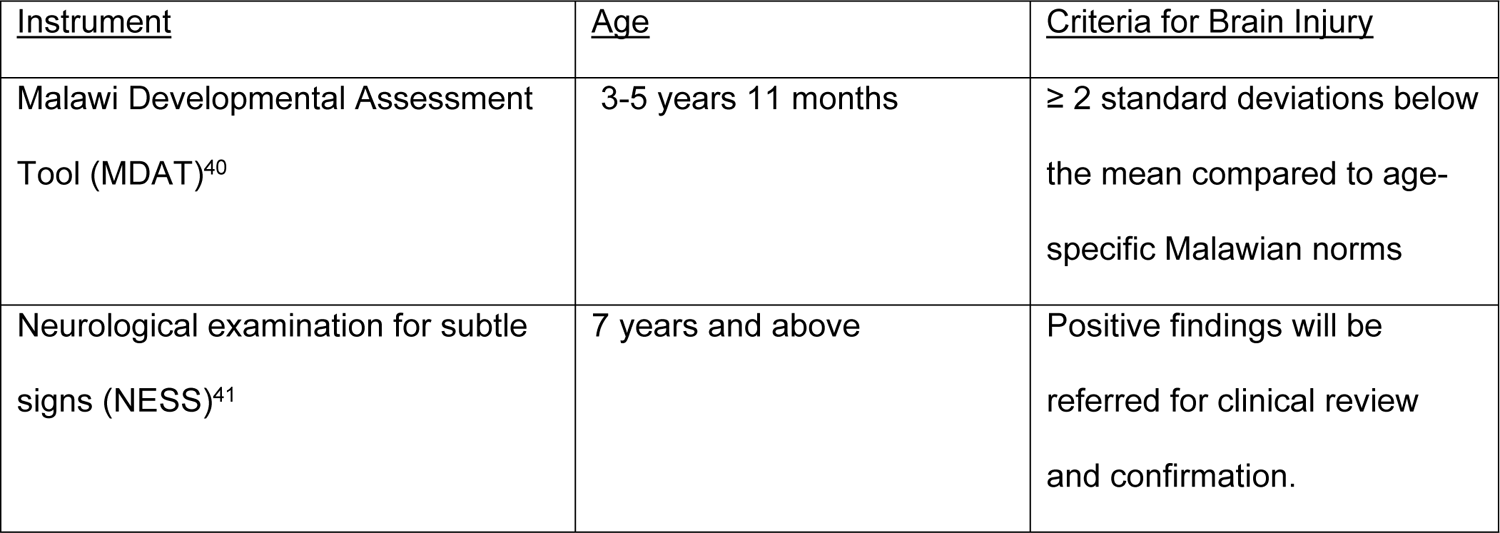
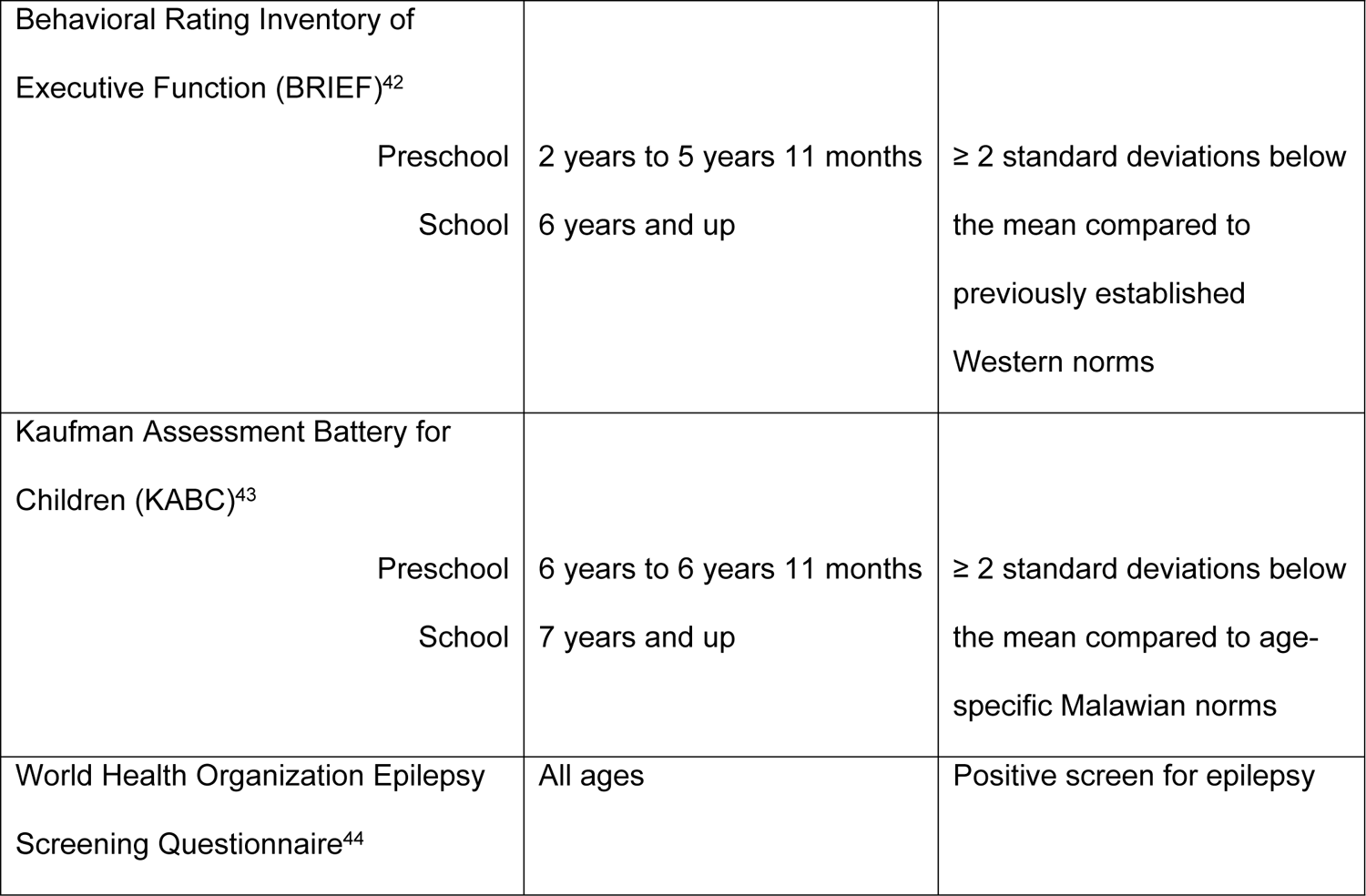
Neurodevelopmental & Behavioral Assessments and Ages for Testing.

| <u>Instrument</u> | <u>Age</u> | <u>Criteria for Brain Injury</u> |
| --- | --- | --- |
| Malawi Developmental Assessment Tool (MDAT)<sup>40</sup> | 3-5 years 11 months | ≥ 2 standard deviations below the mean compared to age-specific Malawian norms |
| Neurological examination for subtle signs (NESS)<sup>41</sup> | 7 years and above | Positive findings will be referred for clinical review and confirmation. |
| Behavioral Rating Inventory of Executive Function (BRIEF)<sup>42</sup> |  |  |
| Preschool | 2 years to 5 years 11 months | ≥ 2 standard deviations below the mean compared to previously established Western norms |
| School | 6 years and up |  |
| Kaufman Assessment Battery for Children (KABC)<sup>43</sup> |  |  |
| Preschool | 6 years to 6 years 11 months | ≥ 2 standard deviations below the mean compared to age-specific Malawian norms |
| School | 7 years and up |  |
| World Health Organization Epilepsy Screening Questionnaire<sup>44</sup> | All ages | Positive screen for epilepsy |

#### Kidney Function

Cr will be determined based upon analysis of whole blood captured via finger prick using a STAT Sensor Creatinine Meter. Urine samples will be obtained for ACR measurements as well. In Zambia, these will be obtained through a commercial lab. In Malawi, these will be assessed with simultaneous measurement of urine albumin and creatinine on a Beckman Coulter AU480 machine with subsequent calculation of the ratio of the two analytes. If any clinically relevant findings are identified, these will be discussed with the parent/guardian and the appropriate referral for care will be made.

### Dissemination

Findings from this work will initially be presented at appropriate local venues before broader dissemination. For example, University Teaching Hospital’s Neurology Research-in-Progress weekly meeting in Lusaka, Zambia; Zambia’s Annual National Health Research Conference in Lusaka, Zambia; the Wellcome Trust’s Cutting-Edge weekly meeting, in Blantyre, Malawi and/or Kamuzu University of Health Science’s Research Dissemination Day in Blantyre, Malawi. With approvals from Zambia’s National Health Research Authority, findings will then be submitted to peer reviewed journal(s) for publication and wider dissemination. Copies of published papers will be submitted to the associated ethical review boards and the Zambian National Health Research Authority.

### Risks to Subjects

Risks include loss of privacy, pain from finger prick, annoyance at the study assessments and adverse effects from sedation. MRI-specific risks are largely associated with implanted devices, which are not in use in the study populations of interest.

The study assessments including the neuroimaging are expected to take less than 2 hours. Regarding sedation risks--in our experience, Zambian and Malawian children as young as 6 years are able to tolerate clinical MRIs without sedation with a parent in the scanning room to comfort and encourage them. In Malawi, an existing sedation protocol utilizing chloral hydrate that has been in place for several years will be used. No serious adverse events have occurred in relation to chloral hydrate use in Malawi to date. An estimated 6% of Malawian children given chloral hydrate develop a paradoxical hypervigilance sometimes with irritability and agitation lasting 4-6 hours. Nausea, vomiting and an allergic reaction are listed side effects of chloral hydrate though these have not been seen during clinical use in children in Malawi over the last decade. Children with a history of an allergy to or adverse reactions to chloral hydrate and those with a history of a paradoxical response to other sedatives will not undergo sedation. Side effects for chloral hydrate are dose related. Although standard protocol for clinical use of chloral hydrate indicates repeated dosing if needed, no repeat dosing will be undertaken in this study. Children who cannot cooperate sufficiently for MRI acquisition and for whom sedation is deemed inappropriate or unsuccessful will not be imaged and their performance on the neurodevelopmental and behavioral assessments will be used to identify brain injury.

In Zambia where chloral hydrate is not available, an alternative sedation strategy will be used--melatonin plus diphenhydramine. Melatonin is the standard sedative used for pediatric electroencephalography in Zambia. Diphenhydramine in doses approved for over-the-counter treatment of cold symptoms in children in Zambia will be added to the melatonin. Should a child experience any adverse effects from sedation, a study clinician will evaluate them immediately. Most anticipated side effects are self-limited and will resolve without intervention. If needed, the clinician will provide analgesics for headache, anti-emetics for nausea, and oral rehydration solution for vomiting.

### Potential Benefits to Subjects

Children identified as having neurologic sequelae or kidney injury based upon the evaluations will be referred for appropriate services that are available free of charge at both study sites.

### Cost of Participation

Participants and their families will not be required to cover any of the costs associated with participation. All study activities and procedures, as well as round trip transportation for the children and their parents/guardians for these follow-up assessments, will be provided free-of-charge.

#### Payment for participation

- In Malawi, for each study visit families will be reimbursed for the roundtrip travel costs for the child and guardian. They will also receive remuneration consistent with published guidelines, which is MWK4500.^45^
- In Lusaka, for each study visit families will be reimbursed for the roundtrip travel costs for the child and guardian. They will also receive a total of ZMW260 for their time lost from work, food during the testing day and their child’s completion of the assessments.
- In Chipata, for each study visit we will provide round trip transportation and overnight accommodation for the child and a parent/guardian to travel to Lusaka. They will also receive ZMW1060 for their time lost from work, food while traveling and the child’s completion of the assessments.

#### Subject withdrawals

Parents/guardians may withdraw their children from the study at any time without impacting the child’s clinical care or any other adverse consequences.

#### Privacy and Confidentiality of Subjects and Research Data

To limit risk of loss of privacy, only the study ID number will identify all laboratory specimens, evaluation forms, reports, and other records that leave the sites. All paper records will be kept in secure study offices in locked filing cabinets. All computer entry and networking programs will be done using study IDs only. Study data using only study IDs will be stored as follows: paper-based in the secure study offices at each site available only to study staff; neurodevelopmental & behavioral outcomes and NeuroInterp data on the University of Rochester’s Research Electronic Data Capture (REDCap) and Malawi Malaria Alert Centre (MAC) server, and neuroimaging files in DropBox with secured folders accessible only to study staff. Study staff will ensure that imaging data and scanned paper-based forms with graphical or pictorial data contain only the study ID and are saved in secured DropBox (i.e., non-public) folders. The study offices are secured rooms with lockable file cabinets. All computers are password-protected.

#### DropBox folders containing de-identified data are non-public

Clinical information will not be released without permission of the participant’s parent or guardian, except as necessary for monitoring by the appropriate institutional review boards (IRBs), the US National Institute of Neurological Disorders and Stroke, the US Office for Human Research Protections, the sponsor, or the sponsor’s designee. If a family needs to be re-contacted for any reason (e.g., an imaging finding that is determined to warrant clinical follow-up), the study ID in this observational study will be cross-referenced against the contact details for each child’s family that are maintained as required in the study pharmacy for the RCT.

#### Possible constraints

This MRI ancillary study will commence after the Fever RCT is underway and as such we anticipate some loss to follow-up even among those who consent for future contact prior to discharge from the Fever RCT study.

#### Data/Sample storage

De-identified imaging and neurodevelopmental & behavioral outcomes will be stored for future use in secured DropBox folders and on the University of Rochester and Blantyre-based Malaria Alert Centre servers.

## Data and Safety Monitoring Plan

This is an observational study. Potential adverse events include breach in confidentiality and any injuries or inconveniences related to participation in the clinical assessments and/or imaging acquisition. Any adverse events identified will be reported to the appropriate institutional review boards. Although not anticipated, should any Serious Adverse Events occur during the conduct of this observational study, the Local Study Monitors serving the RCT will be asked to review with their assessments provided to the overseeing IRBs.

### Data Analysis Plan

The analysis of the presence of a brain injury (yes/no) will involve fitting a logistic regression model with RCT treatment allocation group as the factor of interest, country as a stratification factor, and a stratified propensity score^46^ to account for the fact that the treatment groups will no longer be the original randomized groups due to participants refusing consent and loss-to-follow-up. The propensity score will be estimated using a logistic regression model with treatment group as the outcome variable and baseline covariate information that is thought to be relevant to predicting adverse neurological outcome. These covariates include disease severity (presence/absence of cerebral malaria), seizures prior to admission, admission temperature, admission creatinine, sex, and age. The estimated treatment group odds ratio, along with its associated 95% confidence interval and p-value, will be derived from this model. Similar analyses will be conducted for the other outcomes of interest including the presence of the specific imaging outcomes outlined in Aim 2, the presence of blood products; and the prevalence of chronic kidney disease based upon an ACR of >1.2. For the ordinal gliosis outcomes in Aim 2, a proportional odds logistic regression model will be employed; if the proportional odds assumption appears to be seriously violated, a partial proportional odds logistic regression model or a multinomial logistic regression model will be used instead.

The sample size considerations are based on the logistic regression analyses to be performed in Aim 1. Preliminary data indicate that 34% of pediatric CM survivors in Malawi have abnormal brain MRIs at 1 month or a KABC falling at least 2 standard deviations below the population norms for those over 7 years.^43^ A sample size of 184 subjects (92 per group) will provide > 85% power to detect a reduction in the percentage of subjects with structural injury from 35% in the UC group to 15% in the AA therapy group, using a two-tailed test and a 5% significance level.

### Ethics approval and consent to participate

Written informed consent will be obtained from all participant’s parents or guardians. Study approval will be obtained from the following oversight entities prior to the conduct of this work:

- The University of Zambia’s Biomedical Research Ethics Committee FWA00000338
- The University of Malawi’s College of Medicine Research Ethics Committee FWA00011868
- The University of Rochester’s Research Subjects Review Board (RSRB) FWA00009386
- An intra-institutional reliance agreement will be generated between the University of Rochester’s RSRB and Michigan State University’s Biomedical Institutional Review Board FWA00004556
- The National Health Research Authority in Zambia

No human subjects contact will occur at any of the US-based sites. All data exported to the US will be de-identified prior to transfer. All RSRB determinations regarding this work will be shared with the non-affiliated researchers in a timely fashion.

## Discussion

When completed, this follow-up study of brain injury among children previously in the malaria Fever RCT will determine whether AA treatment in the Fever RCT provided neuroprotection benefits. This study may also help identify adverse events associated with AA treatment including microhemorrhages and kidney injuring leading to CKD.

## Abbreviations

BMPES: Blantyre Malaria Project Epilepsy Study

CNS: central nervous system

CM: Cerebral Malaria

MRI: Magnetic Resonance Imaging

MSU: Michigan State University

RCT: randomized controlled trial.

RSRB: The University of Rochester’s Research Subjects Review Board

Tmax: maximum temperature

PI: Principal Investigator

UNZA BREC: The University of Zambia’s Biomedical Research Ethics Committee

UR: University of Rochester

UNZA BREC: The University of Zambia’s Biomedical Research Ethics Committee

UTH: University Teaching Hospital

## Authors’ contributions

**Conceptualization:** Karl B. Seydel, Gretchen L. Birbeck, Michael Potchen

**Funding acquisition:** Gretchen Birbeck.

**Imaging aspects:** Sam Kampondeni, Michael Potchen, Colleen Hammond

**Investigation:** Gretchen Birbeck, Suzanna Mwanza, Karl B. Seydel

**Methodology:** Gretchen Birbeck, Karl B. Seydel, Michael P. McDermott

**Project administration:** Moses Chilombe, Gretchen L. Birbeck, Suzanna Mwanza

**Resources:** Gretchen L. Birbeck.

**Supervision:** Moses B. Chilombe, Karl B. Seydel, Suzanna Mwanza

**Writing – original draft:** Karl B. Seydel, Gretchen L. Birbeck, Michael Potchen, Michael P. McDermott

**Writing – review & editing:** Moses B. Chilombe, Gretchen Birbeck, Karl B. Seydel, Michael P. McDermott

## Declarations

Moses Chilombe has no disclosures.

Karl B. Seydel has received funding from the US NIH Colleen Hammond has no disclosures.

Suzanna Mwanza has no disclosures

Archana A. Patel has received funding from the US NIH Frank Lungu has no disclosures.

Somwe wa Somwe has no disclosures. Sam Kampondeni has no disclosures.

Michael J. Potchen has received funding from the US NIH and is a paid medical legal consultant.

Michael McDermott has received funding from the US NIH.

Gretchen L. Birbeck has received funding from the US NIH and is a paid consultant for BlueSpark Technologies.

## Data Availability

No datasets were generated or analysed during the current study.

